# The effectiveness of grass, birch, and mugwort pollen allergen immunotherapy over 18 years: A national cohort study in Denmark

**DOI:** 10.1101/2022.11.16.22282380

**Authors:** Peter Bager, Gry Poulsen, Jan Wohlfahrt, Mads Melbye

**Affiliations:** Department of Epidemiology Research, Statens Serum Institut, Copenhagen, Denmark; Aalborg University, Copenhagen, Denmark; Department of Clinical Medicine, University of Copenhagen, Copenhagen, Denmark; Danish Cancer Society Research Center, Copenhagen, Denmark; K.G. Jebsen Center for Genetic Epidemiology, Norwegian University of Science and Technology, Trondheim, Norway; Department of Genetics, Stanford University School of Medicine, Stanford, California, USA

**Keywords:** allergic rhinitis, immunotherapy, observational study, grass pollen, epidemiology, nasal corticosteroids, anti-allergic medication

## Abstract

**Background:** Long-term effectiveness of pollen allergen immune therapy (AIT) for allergic rhinitis (AR) has been studied for different durations. We studied effectiveness over 18 years using nationwide register-data for Denmark.

**Methods:** A register-based cohort study using data on filled prescriptions, 1995-2018, Denmark. In a cohort of 1.1 million intranasal corticosteroid spray users (proxy for AR), we matched users treated with grass, birch or mugwort (GBM) AIT 1:2 with non-treated users on baseline year and 24 characteristics in the 3 years prior to baseline. The primary outcome was the odds ratio (OR) of using anti-allergic nasal spray during the pollen season in the treated vs. non-treated group modelled in a logistic generalized estimating equations analysis using an autoregressive correlation structure and adjusted for years since baseline (one year categories).

**Results:** Among 7,914 AR patients treated with GBM AIT, the OR of using nasal spray 0-5 years after baseline was reduced when compared with 15,520 non-treated AR individuals (0-2 years, odds ratio (OR) 0.81(0.76-0.85); 3-5 years, OR 0.82(0.77-0.87)), but was close to unity or higher thereafter (6-9 years, OR 0.96(0.91-1.03); 10-18 years, OR 1.15(1.06-1.24). In post-hoc analyses, results were more consistent for those who already had 3 of 3 baseline years of use (0-2 years, OR 0.59 (0.54-0.64); 3-5 years, OR 0.64 (0.59-0.71); 6-9 years, 0.82 (0.75-0.91); 10-18 years, 0.95 (0.84-1.08), and in patients using nasal spray in the last pollen season (0-2 years, OR 0.76 (0.72-0.80); 3-5 years 0.84 (0.78-0.91); 6-9 years, OR 0.94 (0.86-1.03); 10-18 years, 0.90 (0.81-1.00)) as opposed to patients who did not use nasal spray in the last pollen season. The post-hoc findings were likewise more consistent for eye drop and oral antihistamine users (secondary outcomes).

**Conclusion:** Patients treated with GBM AIT in routine care to a higher degree stopped using anti-allergic nasal spray 0-5 years after starting the standard three years of therapy. Post-hoc analyses suggested results were more consistent among patients with persistent AR.

## INTRODUCTION

Allergic rhinitis (AR) is a common chronic upper respiratory tract illness, characterized by running nose, sneezing, nasal blockage, as well as itchy and watery eyes. In the majority, these symptoms are caused by IgE-mediated hypersensitivity to plant pollen, and thus triggered in pollen seasons. Allergen-immunotherapy (AIT) with sublingual or subcutaneous exposure to large amounts of the offending pollen allergen is the only treatment with evidence of being disease-modifying,[1-5] and having long-term effects.[3, 6-10] AIT may take months or years to act on symptoms and most AR sufferers rely on faster acting medications such as oral antihistamines, eye drops, and intranasal corticosteroid sprays (INS). Long term effects of AIT are therefore important to study. In clinical trials, longer follow-up is restricted by resources and patient compliance,[11-14] whereas longitudinal observational studies are more feasible and can therefore add unique and valuable information about long term effects of AIT.[3, 6-10, 15-17] Follow-up times up to 15 years after end of therapy have not been studied yet. Using prescription and register data since 1994 for the entire country of Denmark, we therefore studied effectiveness on AR medication use up to 15 years after end of 3 years treatment (18 years of follow-up) in a large, unselected, national patient population treated with GBM AIT, comparing with well-matched controls and also avoiding exclusions e.g. of patients with certain comorbidities or low adherence to therapy.

## METHODS

### Data sources

The study was based on Danish national registers described elsewhere (see STable 1-5). [18-23] Specifically, data was obtained from the Register of Medicinal Product Statistics, which contains daily individual information about filled prescriptions at all pharmacies since 1994 (e.g., AR medication and AIT packages), including filling date, number of dispensed packages, unique Danish product code for each medicine package type, and international Anatomic Therapeutic Chemical (ATC) classification system code (www.medstat.dk/en).[19]

### Ethics

The study was approved by the Danish Data Protection Agency (no. 15/09765). According to Danish law, ethics approval is exempt for such research. Due to the nature of this research, there was no involvement of patients or members of the public in the design or reporting of this study. Direct dissemination to study participants is not possible. Permission from the Danish Health and Medicines Authority has been granted (no. 15/09765).

### GBM AIT treatment

In Denmark, primarily specialized physicians administer pollen AIT to eligible patients with moderate-severe AR, after referral from their general practitioner. While these services are not consistently registered as part of AIT, the AIT prescriptions are. Thus, the allergen extracts/tablets, are prescribed to the patient, who then fills the prescription at a pharmacy (registration) and bring the dispensed AIT packages for the treatment visit. AIT is only partly reimbursed by the otherwise free public health system. Currently, the only three types of pollen AIT marketed in Denmark are for grass (*Phleum pratense*), birch (*Betula verrucosa*), and mugwort (*Artemisia vulgaris*) allergens, mainly prescribed as subcutaneous immunotherapy (SCIT; injections), but for grass also as sublingual immunotherapy (SLIT; melting tablets) since 2007 (STable 1).

### Source population

The combined pollen season for grass, birch, and mugwort (GBM) in Denmark ranged from April until August in the study period.[24, 25] From the entire Danish population, we identified individuals with GBM-induced AR (hereafter denoted “AR individuals” or “INS users”) as individuals >5 years old living in Denmark between 1997 and 2013, who had filled a prescription for intranasal corticosteroid spray (INS, ATC=R01AD) in the GBM pollen season, or two months before, starting 1 February and ending 31 August (hereafter denoted “a pollen season”).

We included INS prescriptions as way to *define* AR individuals, because INS is the most common recommended medication that is specific for AR, while other common AR medications are not likewise specific in the Danish register-based context. For example, the majority of INS requires a prescription from a doctor, while several oral antihistamines are sold over-the-counter and also used for short-term treatment of common cold and influenza symptoms. A small validation study of Danish register-data among children with allergic diseases suggested >1 or >2 INS prescriptions in any year of childhood (mean age 15 years), respectively, has a relatively good sensitivity (84% and 86%) and specificity (66% and 80%), but a lower positive predictive value (PPV) (53% and 65%) for a clinical diagnosis of AR.[26] INS is however also a first-line treatment for chronic rhinosinusitis, perennial allergic rhinitis and non-allergic rhinitis. We conservatively chose to define individuals with GMB-induced AR by INS use during the pollen season, regardless of any use or not outside the pollen season.

### Identification of the GBM AIT treated group

Individuals who were both GBM AIT treated (patients) and had AR (INS users) were identified. To take into account that most patients started GBM AIT in the autumn before next year’s GBM pollen season, we defined an uneven year unit, a baseline sampling year, starting 1 September and ending 31 August. For each baseline sampling year, we included patients with AR with at least one filled prescription of INS in the 3 preceding pollen seasons, and then excluded those who in the 3 preceding sampling years had severe or perennial asthma (defined by hospitalisation/medication), lived outside of Denmark, or had ever received other AITs. We did not exclude INS users if they had INS use both in and outside the 3 preceeding pollen seasons (See Figure 1 and in STable 2 a list of the details of the inclusion and exclusion criteria).

**Figure 1.**
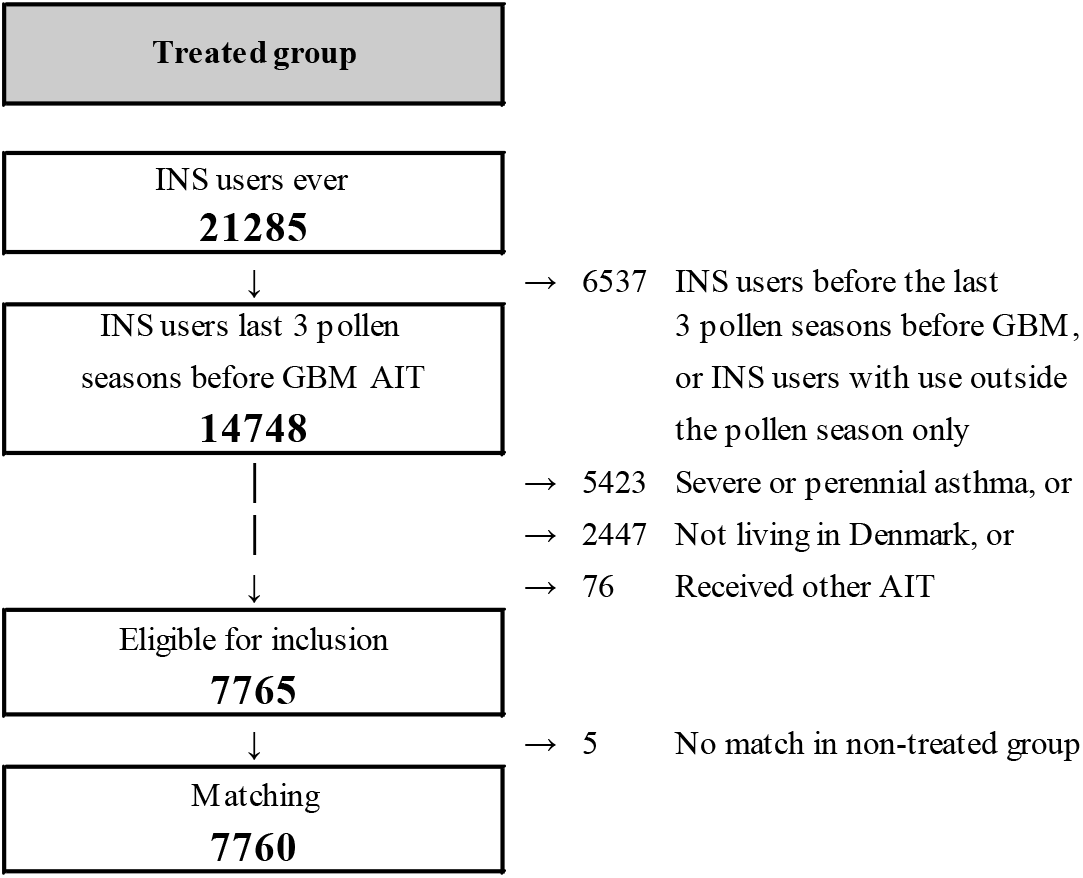
Flowchart for identification of the Grass-Birch-Mugwort (GBM) Allergen Immunotherapy (AIT) treated group, Denmark, 1994-2016 (total data period).

### Identification of the non-treated group (no history of AIT)

Individuals who had no history of receiving AIT and had AR (INS users) were identified for each baseline sampling year according to the same in/exclusion criteria as for the GBM AIT treated group (see STable 3). This was done to allow follow-up of treated and non-treated subjects through the same pollen seasons since baseline.

### Matching

To account for differences in baseline characteristics between GBM AIT treated and non-treated individuals with AR, a propensity score was computed for each baseline sampling year based on socio-demographic characteristics, detailed use of AR medication for the last 3 pollen seasons, family history of asthma and AR, data on medication use and hospitalizations related to non-severe asthma and atopic dermatitis, contacts to own doctor and number of medication groups used (ATC main groups A to V).[27]Propensity scores were computed for a total of 24 characteristics (see STable 4). Both total AR medication use in latest 3 pollen seasons and AR medication use in the immediately preceding pollen season were included in the propensity score model because we after first attempt at matching data found that the GBM AIT group was characterized by a particular high proportion of users of medication in the last pollen season before study entry. Number of contacts to own doctor and number of different ATC main groups of prescription medication used were included with an interaction with age. For each baseline sampling year, each GBM AIT treated patient with AR was matched to two non-treated individuals with AR using optimal matching on the logit of the propensity score (see STable 3). The matched groups were considered well-balanced if the standardized differences were below 10%.[28]The matched individuals were followed from the date of the treated individual’s/patient’s first filled prescription of GBM AIT (date of baseline or “index date”) to 31 August 2016, death, or migration, whichever came first. Individuals from the GBM AIT group were also eligible as non-treated individuals but only in the years before starting on GBM AIT and were thus censored as non-treated individuals the day before they started on GBM AIT on the index date (This was the case for 392 patients)

### Outcome (AR medication users)

The primary outcome was use (vs no use) of <u>at least</u> one nasal spray in the pollen season, and secondary outcomes were similarly defined for use of oral antihistamines (<u>at least</u> one package of tablets) and anti-allergic eye drops (<u>at least</u> one container) (STable 5). There is only one nasal spray or one eye drop container in a prescribed package.

### Statistical analysis

Individuals contributed with information on AR medication for each pollen season during their follow-up. Thus, to take the potential within-subject correlation into account, the odds of using AR medication in GBM AIT treated versus non-treated were modelled as an odds ratio (OR) by a logistic generalized estimating equations analysis using an autoregressive correlation structure and adjusted for years since baseline (one year categories). The main analysis estimated the effectiveness of GBM AIT on each of the three outcomes of AR medication use (with nasal spray use being the primary outcome) within four time intervals since baseline: 0-2, 3-5, 6-9, and 10-18 years. The intervals were chosen because in the 0-2 years interval, up-dosing and maintenance dosing takes place (see STable 1), in the 3-5 years interval a proportion of patients continue dose maintenance, while the 6-9 and 10-18 years intervals would represent the long-term effectiveness of interest, ie. continuing after end of therapy. The Main analysis was performed on the intention-to-treat population, which we defined as all individuals starting treatment (or “exposed population”, ie, those having <u>at least</u> one filled prescription for GBM AIT starting on the index date), and the matched non-treated individuals. The main as well as several additional analysis were performed for the primary outcome nasal spray users and then also for the secondary outcome eye drops and oral antihistamine users. All the additional analysis were performed post-hoc (ie. they were planned after the main analysis), and included effect modification by prebaseline AR medication (1 out of 2, 2 out of 2, and 3 out of 3 prebaseline years of use) and by type of AIT allergen (Grass, Birch, Mugwort, Mixed), which we investigated by introducing interaction terms. Furthermore, another additional analyses conditioned on the use of AR medication last pollen season (yes/no), and were likewise investigated by introducing interaction terms. Lastly, we performed additional analyses focusing on alternative outcomes. For example, to examine bias from different health seeking behaviour by treatment group, we calculated and plotted over time the mean number of different medication groups that individuals used (main ATC anatomical groups) and the mean number of contacts to G.P, by treatment group. To look at use rather than users, we plotted over time the amount of nasal sprays dispensed by treatment group. Finally, to look at asthma as outcome in a final additional analysis, the main analysis was performed with asthma medication as outcome. All analyses were performed with SAS version 9.4 (SAS Institute, Inc., Cary, North Carolina).

## RESULTS

We identified 26,949 new GBM AIT treated patients aged >5 years in Denmark in the period 1.9.1997 to 31.8.2013, of which 19,042 had AR (INS users), and among these 7,760 were eligible for inclusion in the study (4,373 grass, 1,374 birch, 60 mugwort, and 2134 a combination of the 3 AITs). Major reasons for exclusion were not being INS user in the last 3 pollen seasons (55% of all new GBM AIT) or having severe or perennial asthma the last 3 years before baseline (20% of all new GMB AIT) (Figure 1). Each GBM AIT treated individual was successfully matched to two non-treated individuals resulting in a sample of 15,520 non-treated AR individuals. Before matching, the GBM AIT treated group was substantially younger and had a higher use of all types of AR prescription medication than other AR individuals (data not shown). After propensity score matching, the treated and non-treated group were well balanced, with the standardized difference well below 10% for all covariates (Table 1). Assuming one GBM AIT treatment package lasts 6 months, the cumulative percentages of patients ending their last treatment 4, 5, and 6 years after first treatment was 70%, 87%, 94%, respectively (data not shown).

**Table 1.** Baseline characteristics in the propensity-score matched study population, Denmark 1994-2016. See full definitions of variables in STable 4.

| Baseline characteristic |  | Propensity-score matched sample (1:2) |  |  |
| --- | --- | --- | --- | --- |
|  |  | GBM AIT treated | Not treated | Std. diff. |
| **Total number of AR individuals** |  | 7760 | 15520 |  |
| **Baseline sampling years** (dates 1 September to 31 August each year) |  |  |  |  |
|  | 1998-2001 | 20.8% | 20.8% | 0.00 |
|  | 2002-2005 | 27.8% | 27.8% | 0.00 |
|  | 2006-2009 | 22.2% | 22.2% | 0.00 |
|  | 2010-2013 | 29.2% | 29.2% | 0.00 |
| **Demographic characteristics** |  |  |  |  |
| Women |  | 43.9% | 44.3% | -0.01 |
| Age at 1 September in baseline sampling year | 5-9 years | 9.8% | 9.1% | 0.02 |
|  | 10-14 years | 17.9% | 18.5% | -0.01 |
|  | 15-24 years | 20.2% | 20.9% | -0.01 |
|  | 25-34 years | 22.3% | 22.4% | -0.00 |
|  | 35-44 years | 18.2% | 18.4% | -0.01 |
|  | 45+ years | 8.2% | 8.0% | 0.01 |
| Time since first prescription of INS | 0-1 years | 32.5% | 31.9% | 0.01 |
|  | 2-4 years | 31.4% | 31.7% | -0.01 |
|  | 5-9 years | 24.5% | 24.5% | -0.00 |
|  | 10+ years | 11.6% | 11.8% | -0.01 |
| **Use of AR medicine in last three pollen seasons (prebaseline use)** |  |  |  |  |
| Nasal spray, persistence | 1 year out of 3 | 44.9% | 45.3% | -0.01 |
|  | 2 years out of 3 | 30.5% | 30.5% | -0.00 |
|  | 3 years out of 3 | 24.5% | 24.1% | 0.01 |
| Nasal spray, number of packages in last three pollen seasons | 0-1 pkgs. | 34.6% | 35.2% | -0.01 |
|  | 2 pkgs. | 24.3% | 24.8% | -0.01 |
|  | 3-4 pkgs. | 24.4% | 24.1% | 0.00 |
|  | 5+ pkgs. | 16.6% | 15.9% | 0.02 |
| Nasal spray, number of packages in last pollen season | 0 pkgs. | 26.8% | 26.9% | -0.00 |
|  | 1 pkg. | 45.2% | 46.0% | -0.01 |
|  | 2-3 pkgs. | 22.9% | 22.1% | 0.01 |
|  | 4+ pkgs. | 5.2% | 5.0% | 0.01 |

|  |  | Propensity-score matched sample (1:1) |  | Std. diff. |
| --- | --- | --- | --- | --- |
|  |  | GBM AIT treated | Not treated |  |
| Eye drops, number of last three pollen seasons where eye drops were used | 0 years out of 3 | 24.2% | 23.7% | 0.01 |
|  | 1 year out of 3 | 28.7% | 30.0% | -0.02 |
|  | 2 years out of 3 | 22.8% | 22.8% | 0.00 |
|  | 3 years out of 3 | 24.3% | 23.5% | 0.02 |
| Eye drops, number of packages in last three pollen seasons | 0 pkgs. | 24.2% | 23.7% | 0.01 |
|  | 1 pkg. | 22.7% | 23.9% | -0.02 |
|  | 2 pkgs. | 16.3% | 16.6% | -0.01 |
|  | 3-4 pkgs. | 19.1% | 19.2% | -0.00 |
|  | 5+ pkgs. | 17.7% | 16.6% | 0.03 |
| Eye drops, number of packages in last pollen season | 0 pkgs. | 42.7% | 43.5% | -0.01 |
|  | 1 pkg. | 32.4% | 32.6% | -0.00 |
|  | 2 pkgs. | 14.1% | 14.1% | 0.00 |
|  | 3-4 pkgs. | 10.8% | 9.8% | 0.03 |
| Oral antihistamines, number of last three pollen seasons where oral antihistamines were used | 0 years out of 3 | 18.5% | 17.6% | 0.02 |
|  | 1 year out of 3 | 26.8% | 27.5% | -0.01 |
|  | 2 years out of 3 | 25.6% | 26.0% | -0.01 |
|  | 3 years out of 3 | 29.2% | 28.8% | 0.01 |
| Oral antihistamines, number of packages in last three pollen seasons | 0 pkgs. | 18.5% | 17.6% | 0.02 |
|  | 1 pkg. | 16.6% | 17.2% | -0.01 |
|  | 2 pkgs. | 15.5% | 16.3% | -0.02 |
|  | 3-4 pkgs. | 23.5% | 23.4% | 0.00 |
|  | 5+ pkgs. | 26.0% | 25.5% | 0.01 |
| Oral antihistamines, number of packages in last pollen seasons | 0 pkgs. | 34.6% | 34.2% | 0.01 |
|  | 1 pkg. | 29.4% | 30.4% | -0.02 |
|  | 2 pkgs. | 18.8% | 18.6% | 0.00 |
|  | 3-4 pkgs. | 17.2% | 16.8% | 0.01 |
| **Mild/seasonal asthma<sup>a</sup>, atopic dermatitis** |  |  |  |  |
| Mild/seasonal asthma medication, number of last three years where the asthma medication was used | 0 years out 3 | 69.8% | 69.5% | 0.01 |
|  | 1 years out 3 | 19.2% | 19.1% | 0.00 |
|  | 2 years out 3 | 7.4% | 7.7% | -0.01 |
|  | 3 years out of 3 | 3.6% | 3.6% | -0.00 |

(Table 1 continued)
|  |  | Propensity-score matched<br>sample (1:1) |  | Std.<br>diff. |
| --- | --- | --- | --- | --- |
|  |  | GBM<br>AIT<br>treated | Not<br>treated |  |
| Mild/seasonal asthma medication, number of packages in the last three years pollen seasons | 0 pkgs. | 64.9% | 64.9% | 0.00 |
|  | 1 pkg. | 12.2% | 11.9% | 0.01 |
|  | 2 pkgs. | 7.6% | 7.8% | -0.01 |
|  | 3+ pkgs. | 15.2% | 15.4% | -0.00 |
| Mild/seasonal asthma medication, number of packages in last pollen season | 0 pkgs. | 74.9% | 74.8% | 0.00 |
|  | 1 pkg. | 11.3% | 11.6% | -0.01 |
|  | 2+ pkgs. | 13.8% | 13.6% | 0.00 |
| Hospital diagnosis of allergic rhinitis |  | 9.2% | 8.0% | 0.03 |
| Atopic dermatitis (medication and/or hospital contact within last 3 years<sup>b</sup>) |  | 5.5% | 5.4% | 0.00 |
| Parent<sup>c</sup> with allergic rhinitis |  | 56.9% | 57.3% | -0.01 |
| Parent<sup>c</sup> with asthma |  | 36.5% | 37.0% | -0.01 |
| **General health** |  |  |  |  |
| Ever admitted to hospital due to respiratory illness (excluding asthma) |  | 28.1% | 26.6% | 0.03 |
| Average number of different prescriptions drugs over last three years | 0-1 ATC | 13.5% | 13.7% | -0.01 |
|  | 2-3 ATC | 40.8% | 41.6% | -0.01 |
|  | 4-5 ATC | 28.4% | 26.6% | 0.03 |
|  | 6+ ATC | 17.4% | 18.0% | -0.01 |
| Average number of contacts to own doctor over last three years (in tertiles calculated in full sample) | Lowest | 37.2% | 38.0% | -0.01 |
|  |  | 39.6% | 38.9% | 0.01 |
|  | Highest | 23.2% | 23.1% | 0.00 |

(Table 1 continued)
|  |  | Propensity-score matched sample (1:1) |  |  |
| --- | --- | --- | --- | --- |
|  |  | GBM AIT treated | Not treated | Std. diff. |
| **Socio-economic characteristics** |  |  |  |  |
| Education<sup>d</sup> | Unknown | 22.3% | 22.0% | 0.01 |
|  | Short | 21.7% | 21.9% | -0.00 |
|  | Intermediate | 32.2% | 32.7% | -0.01 |
|  | Long | 23.8% | 23.4% | 0.01 |
| Income (in quartiles calculated in full sample) | Lowest | 11.7% | 11.5% | 0.01 |
|  |  | 17.8% | 18.0% | -0.01 |
|  |  | 25.8% | 25.9% | -0.00 |
|  | Highest | 37.2% | 37.2% | -0.00 |

GBM-AIT grass/birch/mugwort allergen immunotherapy.
<sup>a</sup> Defined as those not using medication for severe asthma and not using perennial asthma medications, also see STable 4.<sup>b</sup> Using definition from Henriksen L et al. (J Allergy Clin Immunol. 2015;136(2):360-366).<sup>c</sup> Coding of parent is based on the national CPR registry, Adoption registry, and Medical Birth Registry, and determined in Statistic Denmark's table FTBARN, based on parent-child relation registered shortly after birth in most cases indicate biological parents (see https://www.dst.dk/da/Statistik/dokumentation/Times/fertilitetsdatabasen/far-foed-adop).<sup>d</sup> Coding of education: Short: Primary education or lower secondary; intermediate: Upper secondary, vocational training, some further education not equivalent of a bachelor's degree; long: Further education equivalent of a bachelor's degree or higher

### Main result

For the primary outcome of nasal spray users, Figure 2 shows the proportion who used nasal spray in the GBM AIT treatment and the non-treatment group by year including the 3 years before baseline. Overall, in both groups, the proportion of nasal spray users increased and peaked in the 3 years before baseline, and then dropped immediately after baseline. In the GBM AIT treated group, the proportion of nasal spray users was lower 0-5years after baseline, when compared with no treatment (0-2 years, OR 0.84, 95% CI 0.81-0.88; 3-5 years, OR 0.88, 95% CI 0.84-0.92), but then remained constant while the proportion in the non-treated group dropped, thereby yielding a relative higher proportion of nasal spray users among treated in the later period (6-9 years, OR 1.03, 95% CI 0.97-1.08; 10-18 years, OR 1.18, 95% CI 1.11-1.26) (Figure 2 and Table 2).

**Figure 2:**
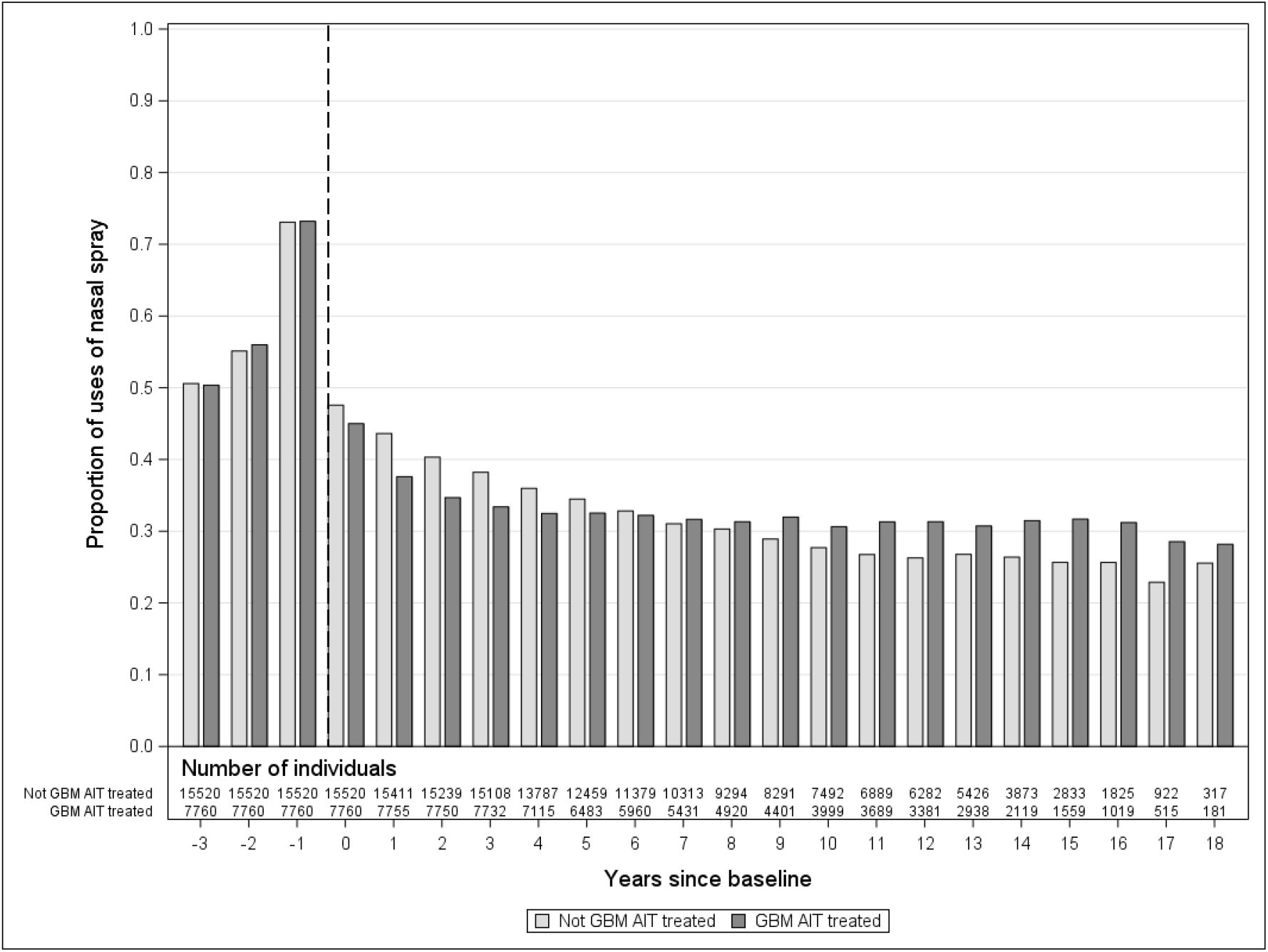
Proportion of users of nasal spray in the pollen season among 7760 GBM AIT treated vs 15520 matched not GBM AIT treated individuals with AR by year since baseline (dotted line), including 3 years before baseline date, Denmark 1994-2016.

**Table 2.** The OR of using AR medication in 7760 GBM AIT treated vs. 15520 matched not GBM AIT treated individuals with AR by years since baseline and type of AR medication, Denmark, 1994-2016.

| Type of AR medication | Years since baseline | GBM AIT treated |  | Not GBM AIT treated |  | OR for using AR medication<sup>b</sup> |
| --- | --- | --- | --- | --- | --- | --- |
|  |  | N<sup>a</sup> | % | N<sup>a</sup> | % |  |
| Nasal spray | 0-2 | 23265 | 39 | 46170 | 44 | 0.84 (0.81, 0.88) |
|  | 3-5 | 21330 | 33 | 41354 | 36 | 0.88 (0.84, 0.92) |
|  | 6-9 | 20712 | 32 | 39277 | 31 | 1.03 (0.97, 1.08) |
|  | 10-18 | 19400 | 31 | 35859 | 27 | 1.18 (1.11, 1.26) |
| Eye drops | 0-2 | 23265 | 35 | 46170 | 37 | 0.93 (0.89, 0.98) |
|  | 3-5 | 21330 | 28 | 41354 | 30 | 0.94 (0.89, 0.98) |
|  | 6-9 | 20712 | 26 | 39277 | 24 | 1.12 (1.05, 1.18) |
|  | 10-18 | 19400 | 24 | 35859 | 19 | 1.27 (1.18, 1.36) |
| Oral antihistamine | 0-2 | 23265 | 50 | 46170 | 45 | 1.24 (1.18, 1.29) |
|  | 3-5 | 21330 | 41 | 41354 | 37 | 1.19 (1.14, 1.25) |
|  | 6-9 | 20712 | 37 | 39277 | 31 | 1.29 (1.23, 1.36) |
|  | 10-18 | 19400 | 34 | 35859 | 26 | 1.44 (1.35, 1.54) |

Abbreviations: GBM AIT grass/birch/mugwort allergen immunotherapy, AR allergic rhinitis, OR odds ratio. <sup>a</sup> Number of observations in time intervals. Since each individual contribute with one observation for each year of follow-up, this number is larger than the number of individuals included in the analysis. <sup>b</sup> The OR for using AR medication is modelled by repeated measure logistic regression using GEE modelling of correlations.

For the secondary outcome of eye drop users, the results of the main analysis was similar (Table 2). However, for secondary outcome of oral antihistamine users, the proportion of users was constantly higher in the GBM AIT group, when compared with the non-treated group (0-2 years, OR 1.24, 95% CI 1.18-1.29; 3-5 years, OR 1.19, 95% CI 1.14-1.25), and even higher after 6 or more years (6-9 years, OR 1.29, 95% CI 1.23-1.36; 10-18 years, OR 1.44, 95% CI 1.35-1.54), despite the two groups being well-matched on prebaseline use (Table 1 and 2).

In an additional analysis, the main result were stratified on years of prebaseline AR medication (see Figure 3 and STable 6). For individuals in the strata with 2 of 3 years prebaseline use of nasal spray (30.5% of GBM AIT treated), the result (see Figure 3) was similar to the main result. For individuals in the strata with 3 of 3 years prebaseline use of nasal spray however (24.5% of GBM AIT treated), the proportion of nasal spray users in the treated group was lower 0-9 years after baseline but then became equal to the proportion in the non-treated group (0-2 years, OR 0.59, 95% CI 0.54-0.64; 3-5 years, OR 0.64, 95% CI 0.59-0.71; 6-9 years, OR 0.82, 95% CI 0.75-0.91; 10-18 years, OR 0.95, 95% CI 0.84-1.08) (Figure 3). For individuals in the strata with 1 of 3 years prebaseline use of nasal spray (44.9% of GBM AIT treated), the proportion of nasal spray users became higher in the GBM AIT treated group already 3 years after baseline. For use of eye drops and oral antihistamine, stratification on prebaseline use yielded a compatible result pattern (STable 6).

**Figure 3:**
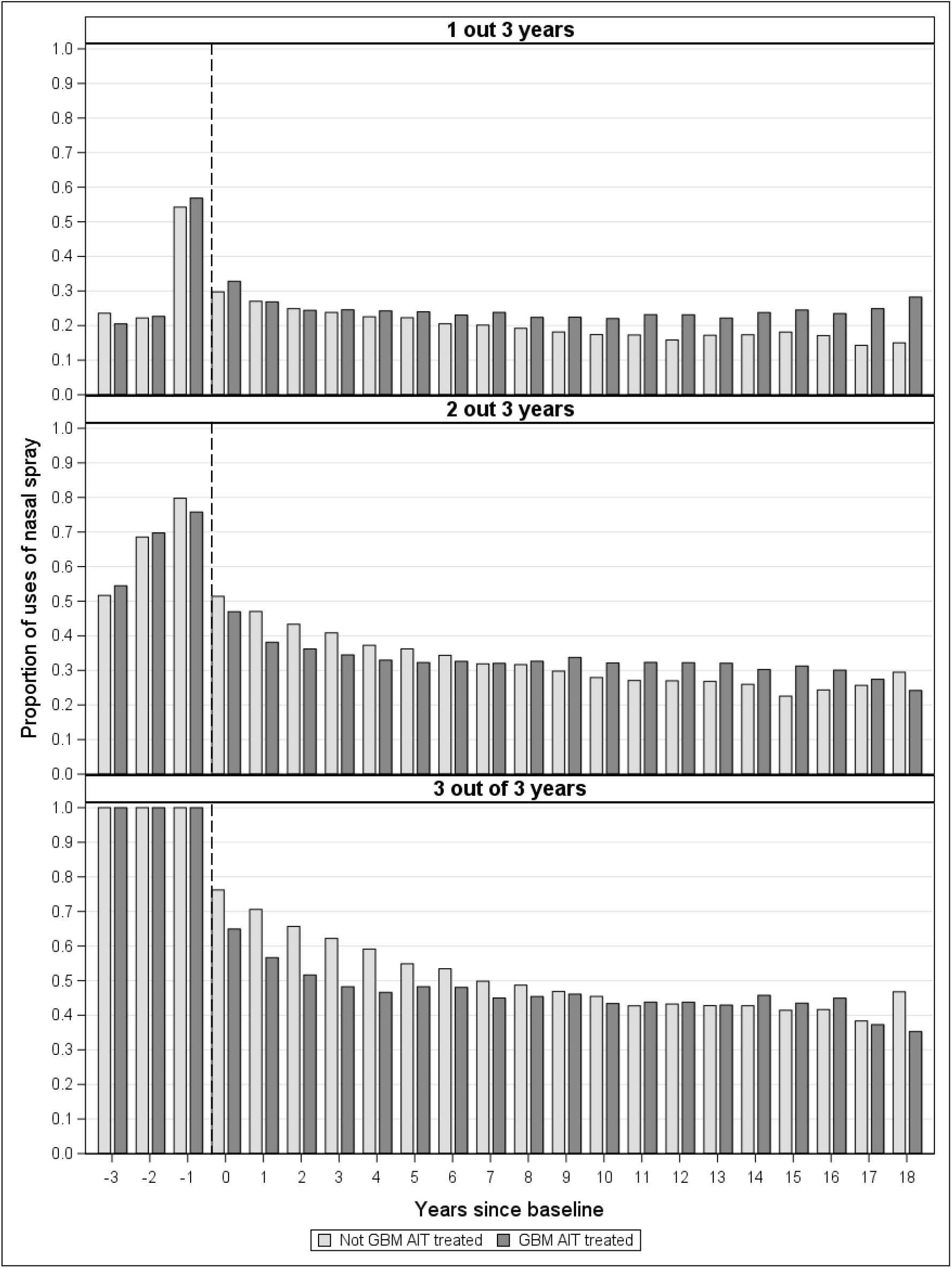
Proportion of users of nasal spray in the pollen season among 7760 GBM AIT treated versus 15520 matched not GBM AIT treated individuals with AR by years since baseline (dotted line) stratified by prebaseline nasal spray use (1, 2, 3 out of 3 years), Denmark 1994-2016

Because our main analyses were based on yearly users of AR medication (and not each individual’s course of use over the years), we also performed an additional analyses, where we estimated the effectiveness of GBM AIT on use of AR medication every follow-up year conditional on an individual’s use of AR medication in the last pollen season (yes/no). Regardless of years since baseline, the proportion of nasal spray users was lower in the treated vs non-treated group within patients who used nasal spray in the last pollen season (0-2 years, OR 0.76, 95% CI 0.72-0.79; 3-5 years 0.86, 95% CI 0.81-0.93; 6-9 years, OR 0.94, 95% CI 0.87-1.02; 10-18 years, 0.94, 95% CI 0.86-1.04), ie. these patients had a tendency to stop using nasal spray, whereas the proportion was higher in the treated vs. non-treated group within patients who did not use nasal spray in the last pollen season (0-2 years, OR 1.02, 95% CI 0.95-1.10; 3-5 years 1.11, 95% CI 1.04-1.19; 6-9 years, OR 1.23, 95% CI 1.15-1.32; 10-18 years, 1.33, 95% CI 1.23-1.44), ie., such patients had a tendency to start using nasal spray when not having used nasal spray in the last pollen season (Table 3). The findings were similar for the proportion of eye drop users, and even for the proportion of oral antihistamine users (Table 3), and even after stratification for prebaseline use (STable 7, shown for nasal spray users only). In the latter analysis, however, every year those with the lowest prebaseline use (1 of 3 years) had a tendency to start using nasal spray, whereas only some years this was case for those with higher prebaseline uses (2 or 3, of 3 years) (Stable 7).

**Table 3.**
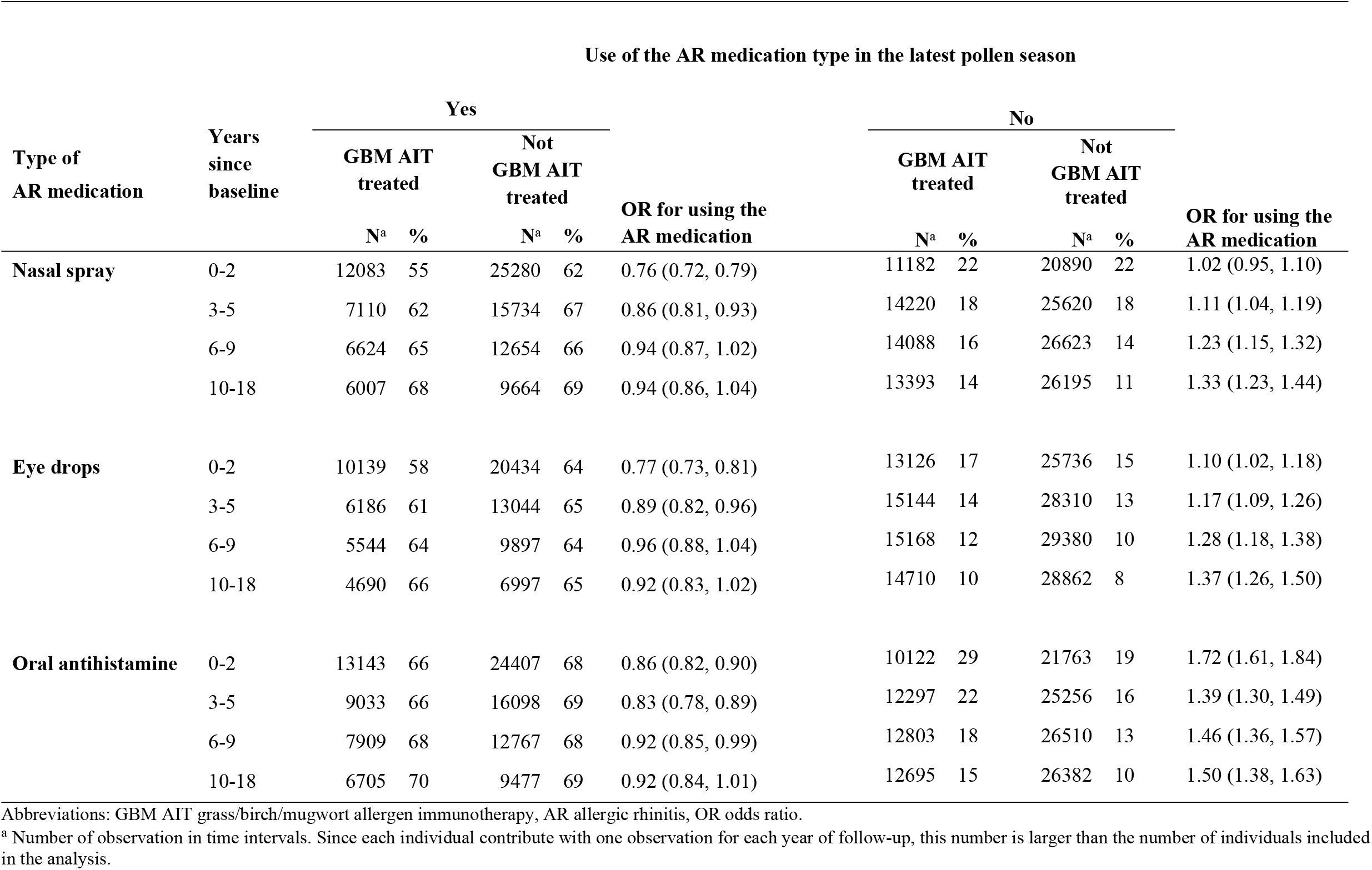
The OR for using nasal spray, eye drops, and oral antihistamine in 7760 GBM AIT treated vs. 15520 matched not GBM AIT treated individuals with AR by years since baseline and use of the AR medication type in the latest pollen season (yes/no), Denmark, 1994-2016.

| Use of the AR medication type in the latest pollen season |  |  |  |  |  |  |  |  |  |  |  |
| --- | --- | --- | --- | --- | --- | --- | --- | --- | --- | --- | --- |
| Type of<br>AR medication | Years<br>since<br>baseline | Yes |  |  |  |  | No |  |  |  |  |
|  |  | GBM AIT<br>treated |  | Not<br>GBM AIT<br>treated |  | OR for using the<br>AR medication | GBM AIT<br>treated |  | Not<br>GBM AIT<br>treated |  | OR for using the<br>AR medication |
|  |  | N<sup>a</sup> | % | N<sup>a</sup> | % |  | N<sup>a</sup> | % | N<sup>a</sup> | % |  |
| Nasal spray | 0-2 | 12083 | 55 | 25280 | 62 | 0.76 (0.72, 0.79) | 11182 | 22 | 20890 | 22 | 1.02 (0.95, 1.10) |
|  | 3-5 | 7110 | 62 | 15734 | 67 | 0.86 (0.81, 0.93) | 14220 | 18 | 25620 | 18 | 1.11 (1.04, 1.19) |
|  | 6-9 | 6624 | 65 | 12654 | 66 | 0.94 (0.87, 1.02) | 14088 | 16 | 26623 | 14 | 1.23 (1.15, 1.32) |
|  | 10-18 | 6007 | 68 | 9664 | 69 | 0.94 (0.86, 1.04) | 13393 | 14 | 26195 | 11 | 1.33 (1.23, 1.44) |
| Eye drops | 0-2 | 10139 | 58 | 20434 | 64 | 0.77 (0.73, 0.81) | 13126 | 17 | 25736 | 15 | 1.10 (1.02, 1.18) |
|  | 3-5 | 6186 | 61 | 13044 | 65 | 0.89 (0.82, 0.96) | 15144 | 14 | 28310 | 13 | 1.17 (1.09, 1.26) |
|  | 6-9 | 5544 | 64 | 9897 | 64 | 0.96 (0.88, 1.04) | 15168 | 12 | 29380 | 10 | 1.28 (1.18, 1.38) |
|  | 10-18 | 4690 | 66 | 6997 | 65 | 0.92 (0.83, 1.02) | 14710 | 10 | 28862 | 8 | 1.37 (1.26, 1.50) |
| Oral antihistamine | 0-2 | 13143 | 66 | 24407 | 68 | 0.86 (0.82, 0.90) | 10122 | 29 | 21763 | 19 | 1.72 (1.61, 1.84) |
|  | 3-5 | 9033 | 66 | 16098 | 69 | 0.83 (0.78, 0.89) | 12297 | 22 | 25256 | 16 | 1.39 (1.30, 1.49) |
|  | 6-9 | 7909 | 68 | 12767 | 68 | 0.92 (0.85, 0.99) | 12803 | 18 | 26510 | 13 | 1.46 (1.36, 1.57) |
|  | 10-18 | 6705 | 70 | 9477 | 69 | 0.92 (0.84, 1.01) | 12695 | 15 | 26382 | 10 | 1.50 (1.38, 1.63) |

Abbreviations: GBM AIT grass/birch/mugwort allergen immunotherapy, AR allergic rhinitis, OR odds ratio.
<sup>a</sup> Number of observation in time intervals. Since each individual contribute with one observation for each year of follow-up, this number is larger than the number of individuals included in the analysis.

In an additional analyses focusing on each AIT allergen, we observed that the main result was similar for patients treated with either grass, birch, or mugwort AIT (STable 8 and 9), or with the recommended 3 years of treatment defined as <u>at least</u> 4 filled prescriptions of GBM AIT in the first 3 years (STable 10). For patients treated with multiple AITs, the proportion of nasal spray users was lower in the first 10 years after baseline and then became equal to the proportion in the non-treated group (STable 8 and 9).

In an alternative outcome analysis, we examined a concern that the GBM AIT might change patients’ general health seeking behaviour more than for the non-treated group, by using health registrations in general as outcomes. We observed an almost equal use of the different ATC main anatomical groups of medications (excluding AIT prescriptions) in the GMB AIT treated and non-treated group every year (SFigure1a). Furthermore, the long term the use of GP services were also almost equal in the two comparison groups, except in the first 0-5 years after baseline when GP services related to AIT however could not be excluded (SFigure 1b). In a final analysis, we depicted number of prescribed AR medication packages as outcome, with the results shown in SFigure 2. Among others, the figure shows that after baseline, less than 50% used <u>at least</u> one nasal spray, and less than 20% used <u>at least</u> two nasal sprays, in a pollen season.

We did a different additional post-hoc analysis, and explored the effectiveness of pollen AIT on “new” medication use for the outcome of severe or perennial asthma (see STable 11, 12, and 13). Here, we assumed conservatively that any medication use for asthma would be “new” because individuals with mediation use for severe or perennial asthma three years prior to baseline were already excluded from the study population. As in the main analysis with AR medication as outcome, we also observed excesses of users for the asthma medication outcome in the treated vs non-treated group over the 18 years (STable 12), and therefore we again stratified by recent use or not: Thus, among patients who used asthma medication last year, the proportion was lower in the treated group (0-2 years, OR 0.80, 95% CI 0.67-0.94; 3-5 years, OR 0.84, 95% CI 0.72-0.98; 6-9 years, OR 0.93, 95% CI 0.81-1.08; 10-18 years, OR 0.88, 95% CI 0.75-1.02), whereas it still higher among patients who did not use asthma medication last year (STable 13).

## DISCUSSION

This longitudinal study of routine care data showed that patients on GBM AIT to a higher degree stopped using nasal spray in the first 0-5 years but not later. In post-hoc analyses, this finding was more consistent among patients with persistent AR.

### Interpretation

The observed effectiveness in reducing nasal spray users 0-5 years after starting pollen AIT is compatible with findings in clinical trials with up to 5 years of follow-up (<300 pollen AIT patients)[11-14] as well as multiple analyses of a regional German health insurance data with 6 years of follow-up (<9000 pollen AIT patients)[7-9] and more recently 9 years of follow-up.[3]

In post-hoc subgroup analysis, we observed GMB AIT effectiveness in the first 0-5 years was greatest and most consistent in those with the most persistent AR medication use at baseline, going back 3 years. This was true both for reductions in nasal sprays, eye drops, and oral antihistamines users, and could suggest the best result of GBM AIT is achieved in patients with the most persistent AR disease. For those with less persistent AR, we however observed a constant excess of users of oral antihistamine, and after 6 years also an excess of nasal spray and eye drop users. These surprising results suggested an additional effect where AR medication use in subgroups of patients may change over the years clearly for other reasons than the immunological effect of GBM AIT. To investigate this further, we performed another post-hoc analysis conditioning in each follow-up year on use or no use in the last pollen season - thereby better capturing changes in use at the individual level over the years. Here, we first observed that for each follow-up year there was a clear tendency to stop nasal spray use but only after a pollen season *with use*. We interpret this as an immunological effect of GBM AIT on AR among those with recent nasal spray use. Secondly we also observed a tendency to start after a pollen season *with no use* - a tendency which in fact fully described the additional effect. One explanation could be that those with no recent use are also patients with less persistent AR at baseline. We therefore cross-tabulated effectiveness by prebaseline use and recent use, and indeed observed the additional effect was largely, although not entirely, restricted to the subgroup of patients with less persistent AR at baseline and a tendency not to use nasal spray the latest pollen season during follow-up (STable 7). However, other explanations may also exist.

The surprising drop in AR medication users immediately after baseline, regardless of pollen AIT, was an unexpected observation. However, pollen AIT is often considered when AR has become chronic, severe, and/or unresponsive to medication. Because this represents a peak in the course of the disease, a subsequent drop in AR medication use would be expected, and also in the control group because of the matching on a similar duration and severity of AR as in the pollen AIT group, as approximated by the last 3 years of medication use.

Our results for pollen AIT effectiveness for 0-5 years are applicable to a selected group of individuals with AR. For example, by our definition about 10% of individuals more than 5 years old in Denmark had AR (ie. INS use in recent pollen seasons), a proportion somewhat lower than reported for any use of INS.[29] Among the 10%, only 2.2% started on GBM AIT. However, the proportion eligible for GBM AIT is likely larger. For example, 75-80% of AR patients are estimated to have moderate-severe AR.[30] Further studies are warranted to estimate if a reduction in disease burden and costs could be achieved by treating more AR patients with AIT.

### Strengths

The study had several strengths. Foremost, the use of real-world data to address long-term effectiveness of pollen AIT over 18 years. In addition, we included detailed individual level data 3 years before baseline. Among others, this allowed us to match pollen AIT patients to non-treated individuals with a similar baseline level of AR medication and history of allergic diseases, thereby strongly reducing potential confounding by a higher baseline severity and persistence of AR in the pollen AIT group.[31]

We acknowledge that AR medication can only be used as proxy for severity and persistence of symptoms. However, since the national register data presented 1.1 million AR individuals (INS users) available for matching, we could match in detail for both number of packages used in the 3 pollen seasons before baseline (a proxy for severity or intensity of AR) and how many of the 3 pollen seasons the medication was used (a proxy for persistence of AR), and this was possible for each specific AR medication. The two comparison groups were also matched on income and educational level, because previous reports suggested AIT patients e.g. have a higher education level.[32, 33] Such factors were not considered in previous real-world studies of AIT effectiveness, all based on German claims databases.[3, 7-9] We chose a design where comparisons groups were evaluated in the same pollen seasons over time, so that pollen exposure would be relatively equal. We had data to perform several sensitivity analysis of the robustness of results. For example, the main results for GBM AIT were robust for grass, birch, or mugwort AIT received alone or in combinations, and even regardless of a higher number of administered GBM AIT packages. Another sensitivity analysis showed that markers of the general health seeking behaviour, ie. medication use and primary care, were at the same level for pollen AIT patients and non-treated individuals during follow-up, except for an excess G.P. contacts the first 0-5 years. The last result is likely related to GBM AIT referrals and prescribing, and is compatible with the German REACT-study finding that subjects treated with AIT were seen more frequently by specialists during the first 3–5 follow-up years, than non-treated in the prescription database.[3, 15, 17]

### Limitations

A limitation of the study was the lack of data on clinical diagnosis of AR and sensitisation status. However, to identify those suffering from AR we restricted our analysis to individuals using INS, which is the classic common effective and specific medication prescribed for AR. Furthermore, we expected the pollen AIT group to have a diagnosis of AR with moderate-to-severe symptoms and confirmed sensitisation status, because it is a prerequisite for the therapy. In the non-treated group, we cannot outrule some misclassification of AR. However, we matched in detail their AR medication use in the 3 prebaseline years (including INS), with that of the pollen AIT patients. This algorithm is more valid than one used in the only Danish validation study so far, which differently defined AR by one year of INS use only, and reported a positive predictive value of 53% for a clinical AR diagnosis.[26]In addition, a bias from misclassification of AR in the non-treated group would tend to overestimate effectiveness, because of less use in the non-treated group. In contrast, we observed effectiveness was reversed or absent in strata with low prebaseline use, where such bias would be largest, but we also observed that effectiveness was better in the strata with the largest prebaseline use where such bias would be smallest (see STable 6). Thus, our results are not consistent with a bias from misclassification of AR in the non-treated group.

Our main outcome measure was the proportion of users of AR medication. We considered this a more appropriate outcome than number of packages used, because, although 100% used nasal sprays before baseline, less than 50% were users in years after baseline (eFigure 2) – thus, use as outcome would not apply to the entire study population, and it would be unclear what patient settings the resulting findings might apply to. Another outcome we considered but did not analyse, was the proportion of more severe users, ie. users of <u>at least</u> 2 packages of AR medication in the pollen season. However, since they constituted less than 20% of the study population, and since comparing them with those using one or no packages would not provide representative or generalisable effects, we did not perform this analysis. A potential limitation was the lack of data on sensitization status for GBM pollen allergens. This means that a grass-AIT patient could be matched correctly to a grass-allergic patient but also incorrectly with a birch-or mugwort-allergic patients, and so forth. However, because we combined the results of the GBM-AIT, and since the choice of AR medication do not differ by type of pollen allergy, this is unlikely to bias the results. In addition, we included only prescriptions filled close to or during Denmark’s GBM pollen season from April to August.[34]

A potential weakness was that filled prescriptions were a proxy for the actual intake of medication. In addition, many antihistamine products in Denmark are not registered by individual as they are sold over-the-counter (although individuals with chronic disease, e.g. AR, are obligated to prescriptions, for reimbursement purposes). For example, for many antihistamine products, 30-40% of users are over-the-counter users as identified purely from Danish sale registrations (without personal identification) (www.medstat.dk/en). In both cases these weaknesses could potentially bias use to be lower than the true use, but to bias our results the lower use needs to be differential between comparison groups. Given the comparison groups were well matched, also on income and educational level, and that we included only medication dispensed within pollen seasons and in the same year in both groups, we considered this was less likely.

Because it has been discussed whether AIT could prevent asthma,[35]we performed a different additional post-hoc analysis using asthma medication as outcome, keeping in mind that individuals with initial severe or perennial asthma to a large degree were already excluded at baseline. As in the previous main analysis with AR medication as outcome, we also observed, surprisingly, an excess of users for the asthma medication outcome in the GBM AIT group over 18 years. However, again, to interpret an immunological effect of GBM AIT in reducing asthma medication use, we stratified by recent use or not, and then observed GBM AIT actually reduced asthma medication users in the first 0-5 years but only in the strata of those with recent use. However, this post-hoc subgroup analysis should be interpreted with caution as our study was not designed to prevention effectiveness. For example, 25-35% of the study population already had used medication for seasonal or mild asthma at inclusion, and the history of clinical asthma symptoms since birth cannot be accounted for completely using register-based data alone, but require a closer clinical follow-up.

## Conclusion

In conclusion, this longitudinal study over 18 years demonstrated that routine care patients who received pollen AIT, compared to non-receivers, had a lower consumption of nasal spray medication in the first 0-5 years after starting pollen AIT. Post-hoc analyses suggested results were more consistent for patients with persistent AR.

## Data Availability

The data are available for research upon reasonable request to the Danish Health and Medicines Authority, Statistics Denmark, and the Research services for the Danish Health Data authority within the framework of the Danish data protection legislation and any required permission from relevant authorities.

## Abbreviations

AIT: allergen immunotherapy;
AR: allergic rhinitis;
GBM: grass-birch-mugwort;
NS: nasal spray;
INS: intranasal corticosteroid spray;
GP: general practitioner.

## Notes

**Source of Funding:** This study was supported by grants from Aase og Ejnar Danielsens Fond (19-10-0208), Dagmar Marshalls Fond, Fonden til Lægevidenskabens Fremme (A.P. Møller og Hustru Chastine McKinney Møller; 18-L-0194), Hartmann Fonden (A33348), and Helsefonden (16-B0320). Dr Bager was supported by a grant from Oak Foundation (OCAY-12-319). The funders had no role in the study design, data collection and analysis, decision to publish, or preparation of the manuscript.

### Competing Interest Statement

The authors have declared no competing interest.

### Funding Statement

This study was supported by grants from Aase og Ejnar Danielsens Fond (19-10-0208), Dagmar Marshalls Fond, Fonden til Lægevidenskabens Fremme (A.P. Møller og Hustru Chastine McKinney Møller 18-L-0194), Hartmann Fonden (A33348), and Helsefonden (16-B0320). Dr Bager was supported by a grant from Oak Foundation (OCAY-12-319). The funders had no role in the study design, data collection and analysis, decision to publish, or preparation of the manuscript.

